# Development of an Integrated Sample Amplification Control for Salivary Point-of-Care Pathogen Testing

**DOI:** 10.1101/2023.10.03.23296477

**Authors:** Navaporn Sritong, Winston Wei Ngo, Karin F. K. Ejendal, Jacqueline C. Linnes

## Abstract

**Background:** The COVID-19 pandemic has led to a rise in point-of-care (POC) and home-based tests, but concerns over usability, accuracy, and effectiveness have arisen. The incorporation of internal amplification controls (IACs), essential control for translational POC diagnostics, could mitigate false-negative and false-positive results due to sample matrix interference or inhibition. Although emerging POC nucleic acid amplification tests (NAATs) for detecting SARS-CoV-2 show impressive analytical sensitivity in the lab, the assessment of clinical accuracy with IACs is often overlooked. In some cases, the IACs were run spatially, complicating assay workflow. Therefore, the multiplex assay for pathogen and IAC is needed.

**Results:** We developed a one-pot duplex reverse transcriptase loop-mediated isothermal amplification (RT-LAMP) assay for saliva samples, a non-invasive and simple collected specimen for POC NAATs. The ORF1ab gene of SARS-CoV-2 was used as a target and a human 18S ribosomal RNA in human saliva was employed as an IAC to ensure clinical reliability of the RT-LAMP assay. The optimized assay could detect SARS-CoV-2 viral particles down to 100 copies/μL of saliva within 30 minutes without RNA extraction. The duplex RT-LAMP for SARS-CoV-2 and IAC is successfully amplified in the same reaction without cross-reactivity. The valid results were easily visualized in triple-line lateral flow immunoassay, in which two lines (flow control and IAC lines) represent valid negative results and three lines (flow control, IAC, and test line) represent valid positive results. This duplex assay demonstrated a clinical sensitivity of 95%, specificity of 100%, and accuracy of 96% in 30 clinical saliva samples.

**Significance:** IACs play a crucial role in ensuring user confidence with respect to the accuracy and reliability of at-home and POC molecular diagnostics. We demonstrated the multiplex capability of SARS-COV-2 and human18S ribosomal RNA RT-LAMP without complicating assay design. This generic platform can be extended in a similar manner to include human18S ribosomal RNA IACs into different clinical sample matrices.

## Introduction

Due to the COVID-19 pandemic, there has been a rapid increase in the development and availability of point-of-care (POC) and home-based nucleic acid amplification tests (NAATs) for respiratory infections. However, this has also given rise to user concerns regarding the accuracy and effectiveness of these tests and has raised questions about the future of at-home diagnostics for other infectious diseases [1]. Without the same level of quality control and assurance as laboratory-based tests, sample quality and the way it is collected and handled can lead to challenges with accuracy, reliability, and reproducibility [2].

According to the US FDA guidelines for *in vitro* diagnostic devices, internal analytical controls (IACs) are among the essential controls mandated for the industry [3]. In the lab, an IAC is commonly run in parallel with NAATs in order to rule out a false-negative result [4] and to qualify the sample collection process as well as the integrity of the amplification enzymes and conditions in a presence of complex matrix [5]. An IAC, sometimes referred to as a sample adequacy control, can contain a synthetic target sequence of different length, a non-target sequence, or a housekeeping gene of human cells to ensure sample is adequately collected and prepared. IACs are also crucial in POC diagnostics to ensure accurate and reliable detection, particularly when performed at locations with limited access to laboratory facilities [6]. The inclusion of an IAC in POC diagnostics can help to reduce the risk of false-negative results, which can have serious implications for the management of infectious diseases due to delayed or inadequate treatment, potentially resulting in disease transmission, increased morbidity, and mortality [7].

While many emerging publications on NAATs for SARS-CoV-2 show remarkable analytical sensitivity in the laboratory setting [8], assessing clinical accuracy with IAC is often overlooked. Incorporating an IAC into POC NAATs can present challenges such as requirements of more complex assay design and higher volume of samples to run the control, which subsequently leads to additional user step. Nevertheless, there are some works that have integrated an IAC on their diagnostic assays as shown in Table 1. Reported IACs were performed in separated reactions from tests for the virus, leading additional user steps, increased cost of reagents, and increased risk of contamination between samples [9–15]. In contrast, a one-pot multiplexed reaction can streamline the assay workflow, decrease the required sample volume, and mitigate the likelihood of cross-contamination among reaction zones, leading to enhanced assay efficacy [16]. While details are not stated, some portable and benchtop commercial SARS-CoV-2 molecular diagnostic tests such as ID NOW™ COVID-19 2.0, Visby Medical Respiratory Health Test, Aptitude Metrix™ COVID-19 test, and Cue’s COVID-19 Diagnostic Test included IAC in their tests as part of result interpretation [17].

**Table 1.** Examples of NAATs for SARS-CoV-2 detection.

| Target gene | IAC target | Sample matrix | Limit of Detection | Clinical sensitivity (%) | Clinical Specificity (%) | Amplification method | Detection method | Reference |
| --- | --- | --- | --- | --- | --- | --- | --- | --- |
| SARS-CoV-2 Nsp3 gene |  | Synthetic RNA | 100 copies/reaction | N/A | N/A | RT-LAMP, WarmStart Colorimetric LAMP 2X Master Mix, Bst 3.0 DNA Polymerase, SuperScript IV Reverse Transcriptase | Leuco crystal violet colorimetric method | Park et al.[48] |
| SARS-CoV-2 ORF1ab, N genes |  | Synthetic RNA | 10-50 copies/reaction | N/A | N/A | RT-LAMP, WarmStart RTX reverse transcriptase, Bst 2.0 DNA polymerase | LFA (Milenia Biotec) | Bhadra et al.[49] |
| SARS-CoV-2 N, E ORF1a genes | Beta-actin gene, run separately | Synthetic RNA | 1-2 copies/μL in dual primer reaction | N/A | N/A | RT-LAMP, WarmStart Colorimetric RT-LAMP 2X Master Mix | Colorimetric assay | Zhang et al.[10] |
| SARS-CoV-2 N gene |  | Nasal swabs | 100 copies/reaction | 97.5 | 99.7 | RT-LAMP, WarmStart 2X Master Mix | Colorimetric assay | Dao Thi et al.[50] |
| SARS-CoV-2 N gene |  | Nasal swabs | 118.6 copies/reaction | 94 | 90 | RT-LAMP, WarmStart Bst 3.0 DNA Polymerase, WarmStartR RT, Q5 High-Fidelity DNA Polymerase | Colorimetric assay | Lu et al.[40] |
| SARS-CoV-2 N, S genes |  | Nasal swabs | 5 copies/μL | 95.4 | 84 | Enhanced recombinase polymerase amplification (eRPA), RNase H, Superscript IV RT | LFA (Milenia Biotec) | Qian et al.<sup>51</sup> |
| SARS-CoV-2 N gene |  | Nasal swabs | 100 copies/reaction | 100 | 98.7 | RT-LAMP, WarmStart Colorimetric RT-LAMP 2X Master Mix | Colorimetric assay | Back et al.[52] |
| SARS-CoV-2 ORF1ab gene |  | Nasal swabs | 20 copies/reaction | 100 | 100 | RT-LAMP, Bst DNA polymerase, AMV reverse transcriptase | Colorimetric assay | Yan et al.[53] |
| SARS-CoV-2 ORF1ab, N genes |  | Nasal swabs | 90 copies/μL | 71.4 | 100 | RT-RPA Basic Kit with T7 Exo-assisted PNA recognition of RPA amplicon | LFA | Zheng at al.[54] |
| SARS-CoV-2 E, M, and N genes |  | Nasal Swabs | 187 copies/reaction | 100% | N/A | triple-target RT-LAMP (ttRT-LAMP) | Colorimetric assay | Zhang et al[55]. |
| SARS-CoV-2 RdRp gene | RNase P gene, run separately | Nasal swabs | 1,526.76 copies/mL | 91.67% | 100% | FastProofTM 30 min-TTR SARS-CoV-2 RT-LAMP Kit | Colorimetric assay | P. Nuchnoi, P. Piromtong, S. Siribal et al.[56] |
| SARS-CoV-2 N, ORF1a genes | Human actin B gene, run separately | Nasal swabs | 25,000 copies/mL | 87.5 | 100 | RT-LAMP, WarmStart Colorimetric RT-LAMP 2X Master Mix | Colorimetric assay | Anahtar et al.[9] |
| SARS-CoV-2 N, E genes | RNase P gene, run separately | Nasal swabs | 10 copies/μL | 95 | 100 | RT-LAMP, WarmStart DNA Polymerase, WarmStart RTx Reverse Transcriptase, LbCas12a | LFA (Milenia Biotec, TwistDx) | Broughton et al.[15] |
| SARS-CoV-2 ORF1ab gene | 18S rRNA, run separately | Nasal swabs | 1 genome copies /μL with image analysis | N/A | N/A | RT-LAMP, GspSSD 2.0 polymerase, AMV-RT | Intercalating agent SYTO82 (nucleic acid stain) | Garneret et al.[11] |
| SARS-CoV-2 N, ORF1a genes | RNase P gene, run separately | Nasal swabs | 5 copies /reaction | 97.4 | 100 | RT-LAMP, WarmStart, Colorimetric WarmStart 2X Master Mix, Bsm polymerase | Colorimetric assay | Papadakis et al.[12] |
| SARS-CoV-2 N, S genes | RNase P gene, run separately | Nasal swabs and saliva | S gene primer: 10 copies/reaction, N gene primer: 25 copies/reaction | *Only show positive samples with 100% agreement |  | RT-LAMP, WarmStart LAMP master mix | End point fluorescence | Yaren et al.[14] |
| SARS-CoV-2 E/S genes |  | saliva | 200 copies/reaction | 98 | 100 | RT-RPA, reverse transcriptase enzyme (EpiScript) | LFA (Milenia Biotec) | Azmi et al.[57] |
| SARS-CoV-2 N, RdRp, ORF1ab genes |  | Saliva | 200 copies /μL saliva | 97 | 100 | RT-LAMP, WarmStart DNA Polymerase, WarmStart RTx Reverse Transcriptase | Colorimetric assay | Davidson et al.[42] |
| SARS-CoV-2 ORF1ab gene |  | Saliva | 1.4 copies/μL | 96.7 | 97.1 | HP-LAMP, WarmStart Colorimetric LAMP 2X Master Mix | Colorimetric assay | Wei et al.[58] |
| SARS-CoV-2 ORF1ab, N genes |  | Saliva | 100-200 copies/reaction | 85 | 100 | RT-LAMP, WarmStart Colorimetric LAMP 2X Master Mix | Colorimetric assay | Lalli[59] et al. |
| SARS-CoV-2 N gene | Proprietary | Nasal swabs | 1.3 copies/μL | 97.4 | 99.1 | RT-LAMP | Results sent to smartphone through app | Cue's COVID-19 Diagnostic Test[60] |
| SARS-CoV-2 N, ORF1ab genes | B2M RNA | Nasal Swabs | 100 copies/swab | 93.2 | 98.9 | PCR | Lateral flow strip on device | Visby Medical Respiratory Health Test[61] |
| SARS-CoV-2 RdRp Gene | Proprietary | Nasal Swab | 500 copies/swab | 93.3 | 98.5 | Isothermal nucleic acid amplification technology | Result displayed by instrument | ID NOW™ COVID-19 2.0[62] |
| SARS-CoV-2 N, ORF-1 genes | Proprietary | Saliva, nasal swab | Swab: 2000 GE/swab<br>Saliva: 20 GE/μL<br>(GE = genome equivalent) | Swab: 96.7<br>Saliva:90.3 | Swab: 99.0<br>Saliva: 99.3 | Isothermal molecular amplification | Result displayed by device | Aptitude Metrix™ COVID-19 Test[63] |
| SARS-CoV-2 ORF1a gene | 18S rRNA, one-pot duplex | Saliva | 100 copies/μL | 95 | 100 | RT-LAMP, 2X WarmStart LAMP master mix | LFA (BioUstar) | This work |

Isothermal amplification reactions, such as reverse transcription loop-mediated isothermal amplification (RT-LAMP) for the detection of SARS-CoV-2 have recently been highlighted due to their key features: rapid detection without the need for sophisticated equipment or specialized training [18], reagent accessibility [19], comparable sensitivity and specificity [20]. With 6 separate primers required, multiplexed RT-LAMP are commonly not considered. However, multiple publications have demonstrated that RT-LAMP assays can be multiplexed to detect multiple targets in a single reaction [21–23]. These can be achieved by including multiple primer sets targeting different regions of the target sequence, as well as incorporating separate probes to differentiate the amplified products.

Choosing an appropriate sample matrix for viral infection diagnosis is essential as it plays a key role in obtaining reliable diagnostic results. Saliva has been reported as an alternative to nasopharyngeal specimens for respiratory virus testing including SARS-CoV-2 [24]. Overall, saliva sampling offers several advantages in terms of simplicity for users. Saliva collection is user-friendly and can be performed without medical personnel, reducing the burden on healthcare professionals. Due to its non-invasive nature, saliva sampling is less intimidating, especially for children and older individuals who may have difficulty with nasal swab collection [25]. Moreover, the time required for specimen collection and the associated cost of using saliva are significantly lower compared to using nasopharyngeal specimens [26].

Despite being easier to collect, salivary components have been demonstrated to hinder RT-LAMP reactions, posing similar challenges to those observed with nasal swab samples. As a result, false-negative results due to assay failure could be observed. To improve clinical accuracy and ease of use and avoiding the drawbacks of a parallel reaction, we have developed a one-pot duplex RT-LAMP assay that uses human 18S ribosomal RNA (18S rRNA) as an IAC in human saliva with the ORF1ab gene of SARS-CoV-2 as the target pathogen. Our optimized assay can detect SARS-CoV-2 viral particles as low as 100 copies/µL of saliva within 30 minutes. The duplex RT-LAMP for SARS-CoV-2 and IAC can be amplified in one-pot reactions without cross-reactivity, and valid results are easily visualized in triple-line lateral flow immunoassays (LFIAs). The appearance of flow control and IAC lines represent valid negative results, and the flow control, IAC, and test lines represent valid positive results. The duplex RT-LAMP assay was validated directly on clinical saliva samples without prior RNA extraction. The IAC developed here meet the FDA guidelines for *In Vitro* Diagnostic Devices [6] to bring clinically relevant molecular diagnostic into POC settings without complicating platform design.

## 2. Materials and methods

### 2.1 Reagents

Reagents for RT-LAMP reactions included WarmStart® Multi-Purpose LAMP/RT-LAMP 2X Master Mix with UDG from NEB (Ipswich, MA), EvaGreen from VWR International (Radnor, PA), ROX from Thermo Fisher Scientific (Waltham, MA), and nuclease-free water from Invitrogen (Waltham, MA). Pooled human saliva used in assay development was purchased from Innovative Research (Novi, MI). BtsYI and Ddel restriction enzymes were purchased from NEB (Ipswich, MA). The viral templates obtained from BEI Resources (Manassas, VA) included Heat inactivated Novel Coronavirus, 2019-nCoV/USA-WA1/2020, NR-52286 (SARS-CoV-2); Middle East Respiratory Syndrome Coronavirus (MERS CoV), EMC/2012, Irradiated Infected Cell Lysate, NR-50549; and SARS Coronavirus (SARS), NR-9547; and purified genomic RNA from dengue virus (DENV) type 1. All primers, including those conjugated to FITC, biotin and DIG, were ordered from Integrated DNA Technologies (Coralville, IA).

### 2.2 Singleplex RT-LAMP for SARS-CoV-2 detection

The primer set targeting ORF1ab region of SARS-CoV-2 (GenBank: OQ691200.1) was used (Table S1.). The RT-LAMP reactions were carried out by using 2x WarmStart® Multi-Purpose LAMP/RT-LAMP Master Mix in accordance with the New England Biolab standard and 10X Primer Mix containing all 6 LAMP primers (final concentration of 1.6 µM for forward inner primer (FIP) and backward inner primer (BIP), 0.4 µM for forward loop primer (LF) and backward loop primer (LB), and 0.2 µM for forward outer primer (F3) and backward outer primer (B3). The 5’ end of LF and LB were labeled with fluorescein (FITC) or biotin, respectively for LFIA detection. The robustness of primer sets in various saliva percentages (0-30%) were evaluated to determine volume of saliva sample used in the assay. Various concentrations of SARS-CoV-2 viral particles were spiked into saliva samples at concentrations ranging from 0 to 5000 SARS-CoV-2 viral copies/µL to determine the limit of detection (LOD) of the assay. The saliva without viral particles was used as a no template control (NTC). Five (5) µL of saliva sample and 20 µL of mastermix were incubated at 65°C for 30 minutes in a QuantStudio 5 Real-Time PCR machine (ThermoFisher, Waltham, MA). The specificity of optimized RT-LAMP against other coronaviruses was performed by using SARS, MERS CoV, DENV viruses as targets. To validate the amplification process, real-time fluorescence data of EvaGreen intercalating dye and ROX reference dye were recorded. The RT-LAMP amplicons were visualized via LFIA and confirmed via gel electrophoresis using a 2% agarose gel run at 100 V for 50 minutes, stained with ethidium bromide, and imaged using an ultraviolet light gel imaging system (c400, Azure Biosystems, Dublin, CA).

### 2.3 Singleplex RT-LAMP for human RNA in saliva for amplification control

To verify the proper collection of saliva and avoid potential false-negative results due to technical errors, we searched the literature for primer sets targeting ubiquitously expressed genes in human samples. In this work, the primer set targeting human 18S rRNA (GenBank: AL592188.60) developed by Garneret et al [11] was selected (Table S1) since it resulted in consistent results. To confirm that IAC primers are orthogonal to SARS-CoV-2 primers and RNA, *in-silico* PCR validation was carried out using free software − UCSC *In-Silico* PCR [27]. The RT-LAMP of IAC was performed as described in singleplex RT-LAMP for SARS-CoV-2 detection experiment. SARS-CoV-2 spiked water, saliva sample, and total human RNA control (Applied Biosystems, Waltham, MA) were used as a template for RT-LAMP of 18S rRNA. The 5’ end of LF and LB were tagged with digoxigenin (DIG) and biotin, respectively for LFIA detection. The amplification was confirmed by gel electrophoresis and the LOD was determined via LFIA. In the LOD experiment, the total human RNA control was 10-fold diluted from 4.4 x 10^6^ copies/µL and used as templates.

### 2.4 One-pot duplex RT-LAMP of SARS-CoV-2 and human RNA

The optimized duplex RT-LAMP consisted of the two sets of 10x Primer Mix targeting the ORF1ab gene or 18S rRNA in which concentrations of FIP and BIP from both sets were adjusted to 1.0 µM while the concentrations of F3, B3, LF, and LB from both sets remained the same as in the singleplex RT-LAMP conditions. Templates used in these experiments were one target and two target templates. The one target template included SARS-CoV-2 spiked water and saliva without viral particles, while two target template was SARS-CoV-2 spiked saliva. The nuclease-free water was used as NTC in this experiment. The amplifications of duplex RT-LAMP were visualized on triple-line (FITC, DIG, flow control) LFIA strips. The nuclease-free water was used as the NTC to ensure that there is no contamination or cross-reactivity between two primer sets. To assess the LOD of the duplex RT-LAMP assay, saliva samples were spiked with different concentrations of SARS-CoV-2 viral particles ranging from 0 to 5000 SARS-CoV-2 viral copies/µL. The duplex RT-LAMP reactions were prepared in total volume of 50 µL with 5 µL of sample and incubated at 65°C for 30 minutes in the QuantStudio5 (Applied biosystems, Waltham, MA).

### 2.5 Restriction enzyme digestion of duplex RT-LAMP products

To validate the expected amplification products, the duplex RT-LAMP products were digested with the restriction enzyme BtsYI and Ddel that are specific to the products of SARS-CoV-2 and IAC primers, respectively. To determine restriction enzymes that specifically cut the product of SARS-CoV-2 or 18S rRNA RT-LAMP, and not both, the NEBcutter software was used [28]. The templates used in this experiment included SARS-CoV-2 spiked water as SARS-CoV-2 primer product, SARS-CoV-2 free-saliva as IAC primer product, and SARS-CoV-2 spiked saliva as duplex product. The reaction consisted of 5 µL of amplicons, 2.5 µL CutSmart Buffer (NEB, Ipswich, MA), and 1 µL of restriction enzyme. The nuclease-free water was added to fill up reaction volume to 25 µL. The reactions were then incubated at 37°C in water bath for 20-30 minutes. The restriction fragments of duplex RT-LAMP products were visualized using ethidium bromide in a 2% agarose gel.

### 2.6 Clinical sample validation

The developed assay was validated against frozen clinical saliva samples received from Indiana Biobank (Bloomington, IN). The saliva samples were collected in 2 mL cryovials and stored in -80°C upon arrival in our lab. These samples were assigned as study ID 1-30, aliquoted, and used as templates for the standard RT-qPCR and duplex RT-LAMP. Of 30 clinical samples, three (3) SARS-CoV-2 negative samples had insufficient volume to extract RNA and run both RT-qPCR and duplex RT-LAMP. Therefore, additional 3 samples were collected from subjects who were negative for COVID-19 by using RNAPro•SAL™ (Oasis Diagnostics®, Vancouver, WA) in accordance with Purdue University IRB protocol # IRB-2020-968. All clinical samples were heat-inactivated at 95°C for 5 minutes to ensure safety of working conditions in a BSL-2 laboratory [29] and stored at −80°C until use. To validate duplex RT-LAMP against clinical samples, 5 or 10 µL of heat-inactivated samples were used as templates with 45 or 40 µL of RT-LAMP master mix as described in one-pot duplex RT-LAMP experiment. Each sample was run in triplicate and visualized on trip-line LFIAs. The samples for RT-qPCR were extracted by using the QIAamp Viral RNA Mini Kit (Qiagen, Hilden, Germany) according to manufacturer’s protocol. Ten (10) µL of extracted samples was used for RT-qPCR analysis using FDA authorized 2019-nCoV: Real-Time Fluorescent RT-PCR kit (BGI, Shenzhen, China) targeted the ORF1ab gene of SARS-CoV-2 genome and human β-actin gene as the IAC. The RT-qPCR was run twice for each sample. The clinical sensitivity and specificity of duplex RT-LAMP assay were evaluated as followed [30]: Sensitivity = (true positive)/(true positive + false negative); Specificity = (true negative)/(true negative + false positive); Accuracy = (true positive + true negative)/(true positive + true negative + false positive + false negative).

### 2.7 LFIA quantification, statistical analysis, and graphical abstract

The LFIA tests were run in triplicates for each experiment. The LFIA quantification and statistical analysis were performed as described by Phillips et al [31]. Briefly, after 15 mins of initial sample addition, the LFIAs were scanned using Epson V850 Pro Scanner. A custom MATLAB script was used to quantify the test band, which averages the grey-scale pixel intensity of the test band and subtracts the average background pixel intensity 40 pixels below the test band. The LOD was determined by a one-way ANOVA with Dunnett’s *post hoc* test with multiple comparisons using GraphPad prism (GraphPad Software, Boston, MA) of the LFIA test bands of each concentration against the test bands from negative controls (no template) with a 95% confidence interval. The graphical abstract is created in Biorender (Toronto, Canada).

## 3. Results and discussion

### 3.1 Analytical sensitivity and specificity of SARS-CoV-2 RT-LAMP assay in saliva samples

Throughout the stages of infection, the viral load can be as low 10^3^ to 10^5^ copies/mL of saliva sample (equivalent to 1 to 100 copies/µL of saliva) in early stage and spiked to 10^8^ copies/mL (10^5^ copies/µL) in later stage [32]. To evaluate the analytical sensitivity of SARS-CoV-2 RT-LAMP assay, saliva spiked with various concentration of inactivated SARS-CoV-2 particles was used as templates. The time to detection of SARS-CoV-2 in the RT-LAMP assay was displayed as the cycle threshold (Ct) value against various concentrations of SARS-CoV-2. The singleplex RT-LAMP targeting ORF1ab gene was able to detect as few as 100 SARS-CoV-2 viral copies/µL saliva in less than 30 minutes (Figure 1A). The SARS-CoV-2 RT-LAMP products visualized by gel electrophoresis were in a ladder-like pattern (Figure 1B), indicating the successful production of the different length concatemers. The conjugation of the backward loop primer to biotin and the forward loop primer to FITC allowed the result to be readout on LFIAs. The test line of saliva samples with SARS-CoV-2 concentration of 100, 500, and 5000 viral copies/µL were observed (Figure 1C). The presence of the flow control line in all samples suggested that the flow of samples on LFIAs was effective. The analytical sensitivity or LOD of the assay was determined by quantifying test band intensity of each concentration. The custom MATLAB script calculates the average gray-scale pixel intensity of the test band and then subtracts the average background pixel intensity located 40 pixels below the test band. As shown in Figure 1D, there is a statistically significant difference in test band intensity of RT-LAMP products from 100-5000 viral copies/µL in saliva as compared to no template control when using a one-way ANOVA with Dunnett’s *post hoc*. The analysis of gel electrophoresis and LFIAs suggested that our SARS-CoV-2 RT-LAMP assay exhibit the LOD of 100 viral copies/µL saliva, which is in the range of clinically relevant LOD [32]. Moreover, the developed RT-LAMP assay was specific to SARS-CoV-2 RNA and demonstrated no amplified products in the gel nor test band on LFIAs when viral particles from MERS, DENV1, or CoV were used as a template (Figure S1). Taken together, the RT-LAMP assay in saliva is both sensitive and specific for SARS-coV-2 detection.

**Figure 1.**
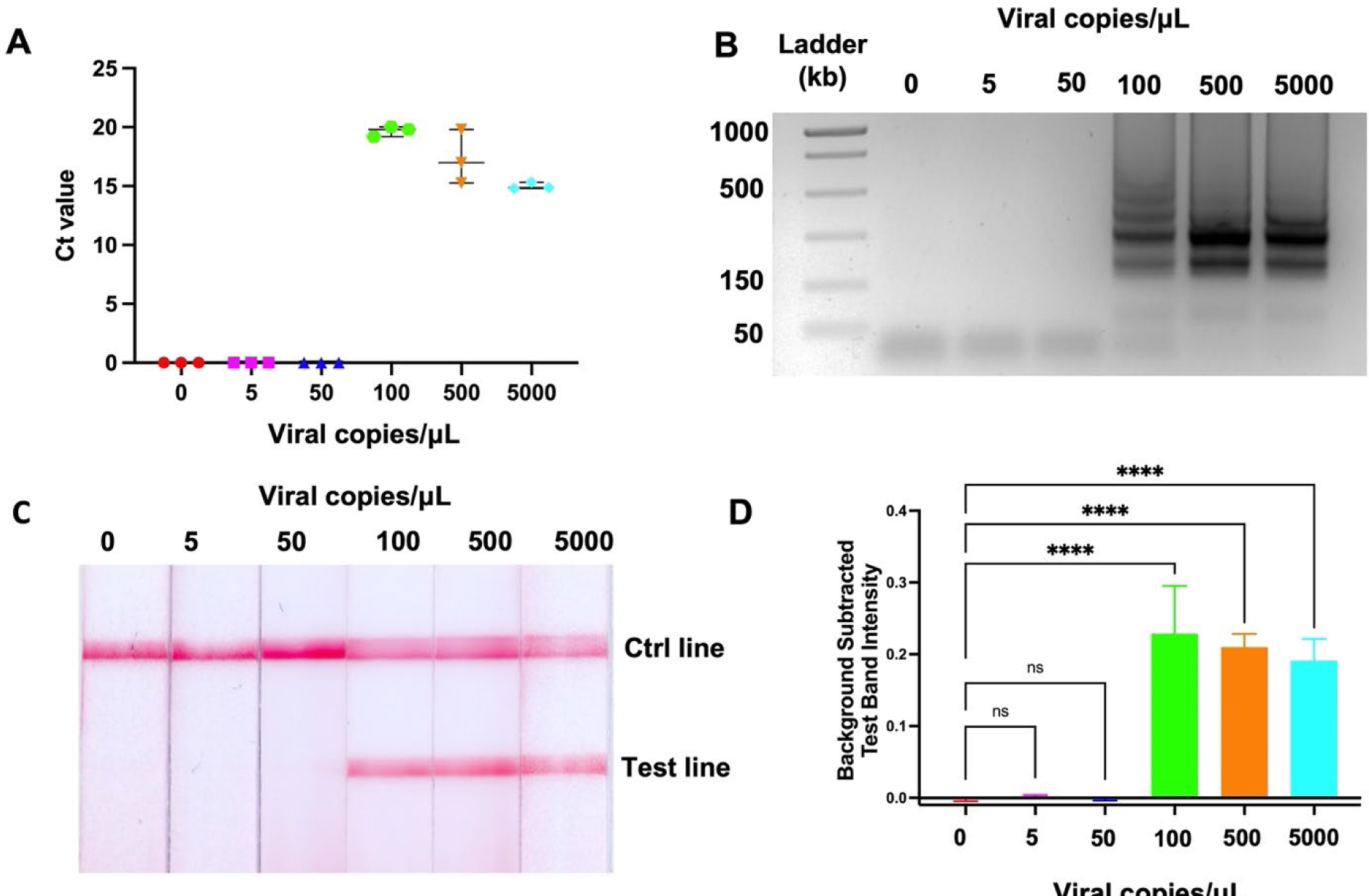
Analytical sensitivity of SARS-CoV-2 RT-LAMP assay in saliva. Ct value of SARS-CoV-2 RT-LAMP in various concentrations (A). RT-LAMP based detection of inactivated viral particles as visualized on gel electrophoresis (B) LFIA (C), and corresponding test band intensity analysis (D). n = 3; **** indicates *p* value ≤ 0.0001. The ladder-like bands on gel electrophoresis indicating successful amplification.

### 3.2 RT-LAMP of 18S ribosomal RNA IAC specific to human RNA in saliva

IACs are one of the required controls according to the US FDA guideline for *in vitro* diagnostic devices for various infectious diseases provided to the industry [3]. Given that saliva is the selected sample matrix for the study, we screened for human genes that are expected to be ubiquitously expressed in human saliva to use as the IAC target. Regardless of the presence of target pathogen, the IAC should be detectable in all samples. Another criterion for IAC in this work is that the IAC primers and target should not cross-react with SARS-CoV-2 primers, or the SAR-CoV-2 target. No matches were found when conducting an *in-silico* analysis of PCR using primers designed for the 18S rRNA against the SARS-CoV-2 genome and primers (Figure S2). This result suggested that IAC primers are orthogonal to SARS-CoV-2 primers and RNA. The amplification plot of 18S rRNA RT-LAMP in Figure 2A demonstrates that sigmoidal curve of the fluorescence signal showed up only when the positive control — total human RNA control and saliva were used as the templates. The amplicons were subsequently analyzed by the gel electrophoresis. As seen in Figure 2B, the gel image displayed the ladder-like bandings when total human RNA control and saliva were used as templates. In contrast, no products were seen on the gel when the template was SARS-CoV-2 particles without saliva. This result confirms that 18S rRNA primers were specific to human RNA control and saliva sample and there is no cross-reactivity against SARS-CoV-2. Therefore, the primer set targeting human 18S rRNA was chosen and incorporated in duplex RT-LAMP as the IAC. It is worth noting that the amount of human 18S rRNA in human saliva can vary depending on several factors including collection method used and the time of day the sample was collected. We ran the LOD of 18S rRNA RT-LAMP and found that the LOD of the assay was 4,400 copies/µL (Figure S3), which is in the range of reported concentrations of 18S rRNA in human saliva by other groups [33].

**Figure 2.**
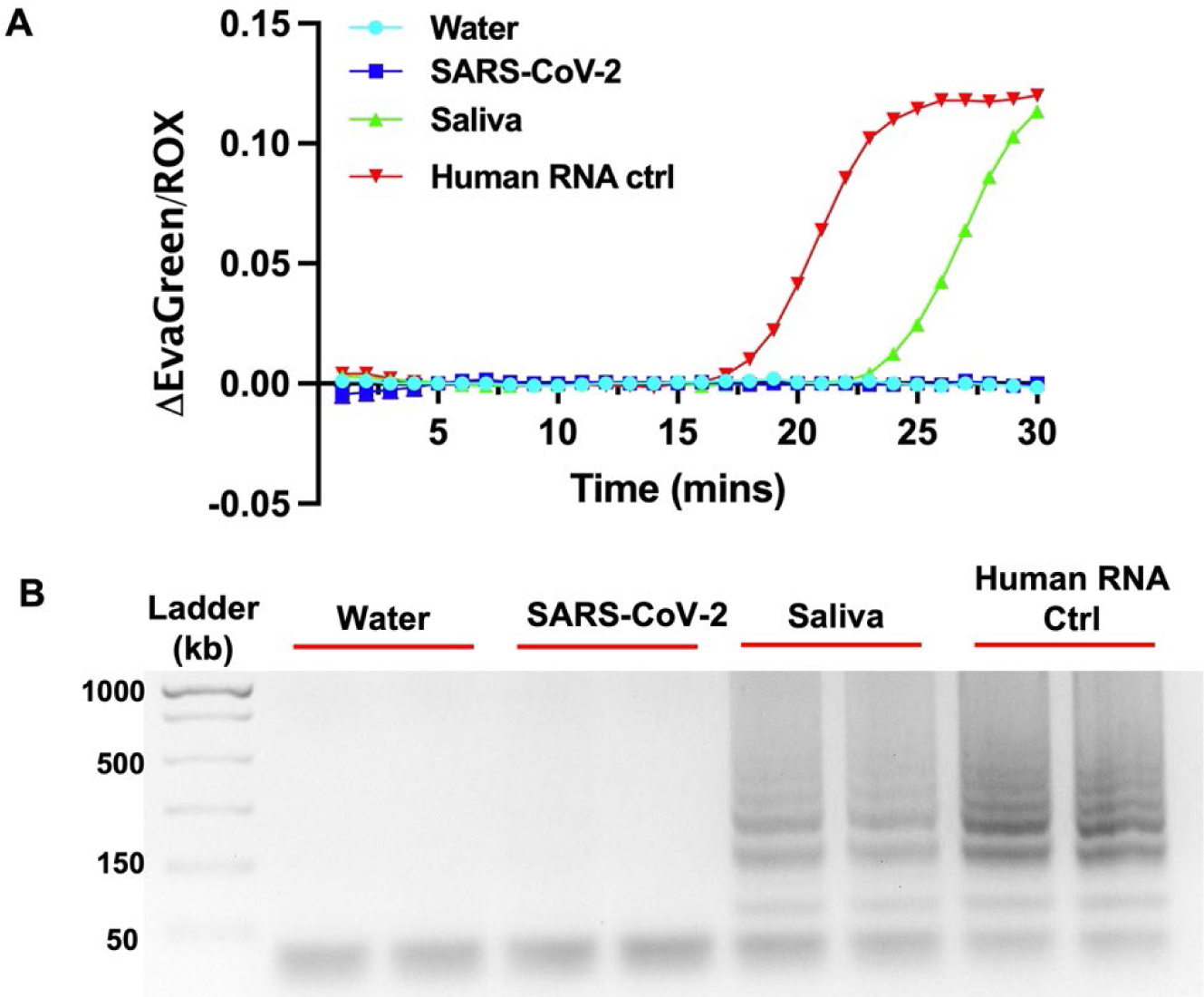
Specificity of IAC primer. Amplification plot (A) and gel electrophoresis (B) of human 18S rRNA RT-LAMP using different templates. The ladder-like bands indicating successful amplification are only present when saliva and human RNA were used as templates. N = 2.

In addition to SARS-CoV-2, saliva could serve as specimen for other respiratory viruses [34] such as Influenza viruses, Respiratory Syncytial Virus (RSV), Adenovirus, and Rhinovirus and even bacterial pathogens such as Group A streptococci causing strep throat, and *Bordetella pertussis* causing whooping cough [35]. This study suggests that the 18S rRNA could be a suitable IAC for other diagnostic tests that rely on saliva samples. Moreover, this knowledge could also be similarly employed for integrated IACs in other clinical sample matrixes such as urine, nasal swabs, and blood. Kretschmer-Kazemi Far et al reported that 18S rRNA is the most abundant RNA species in urine samples [36] and Garneret et al. reported the use of 18S rRNA as the IAC for nasal swabs [11]. By performing 18S rRNA RT-LAMP using whole blood (five (5) µL of undiluted blood), we also demonstrated that 18S rRNA was detectable in freshly collected blood samples (Figure S4.)

### 3.3 Optimization of one-pot duplex RT-LAMP of SARS-CoV-2 and IAC

In general, a one-pot multiplexed reaction can simplify the assay workflow and reduce the amount of sample and reagents required. It can also minimize the risk of cross-contamination between reaction zones and improve the overall assay efficiency. However, a one-pot multiplexed reaction can be more challenging to optimize, and the different targets may compete for limited resources, such as enzymes or primers [37]. Therefore, the choices of primers, concentrations, and assay control template are critical to the optimization process. The duplex RT-LAMP of SARS-CoV-2 and IAC consisting of 2 sets targeting the ORF1ab gene and 18S rRNA was performed in a single tube “one-pot reaction”. Different template setups were used to examine the interactions between additional primer sets and their cross-reactivities. The successful amplification of sample containing one target (SARS-CoV-2 spiked water (W+) and saliva without viral particles (S-)) and two targets (SARS-CoV-2 spiked saliva (S+) are shown on the amplification plot (Figure 3A) and gel electrophoresis (Figure 3B). The NTC showed no banding on the gel, confirming that there was no non-specific amplification or primer-dimers. As seen in Figure 3A, the presence of saliva in SARS-CoV-2 spiked saliva sample (S+) slowed down the amplification of SARS-CoV-2 primers as compared to SARS-CoV-2 in water (W+). This is likely due to factors in the complex matrices such as saliva that interfere with the activity of the enzymes, degrade or inhibit the RNA template or primers, or increase the viscosity of the reaction mixture [38]. The delayed amplification is not specific to the multiplex RT-LAMP primer set used here and has been observed before when complex matrices were introduced into LAMP reaction [39]. The amplicons from different conditions were visualized on a gel were added to LFIAs (Figure 3C). Without templates of both primer sets (NTC) on the W-strip, only the flow control line was present. As anticipated, the W+ strip showed the flow control and test lines, while the S-strip displayed the flow control and IAC lines (Figure 3C). The amplicons of both primers were present on the S+ strip, indicating the successful duplexed RT-LAMP of the target pathogen and IAC in one-pot reaction (n=3). This duplex reaction demonstrates the multiplexing capabilities of 18S rRNA and the ORF1ab gene of SARS-CoV-2 in human saliva in the one-pot reaction platform (Figure 3).

**Figure 3.**
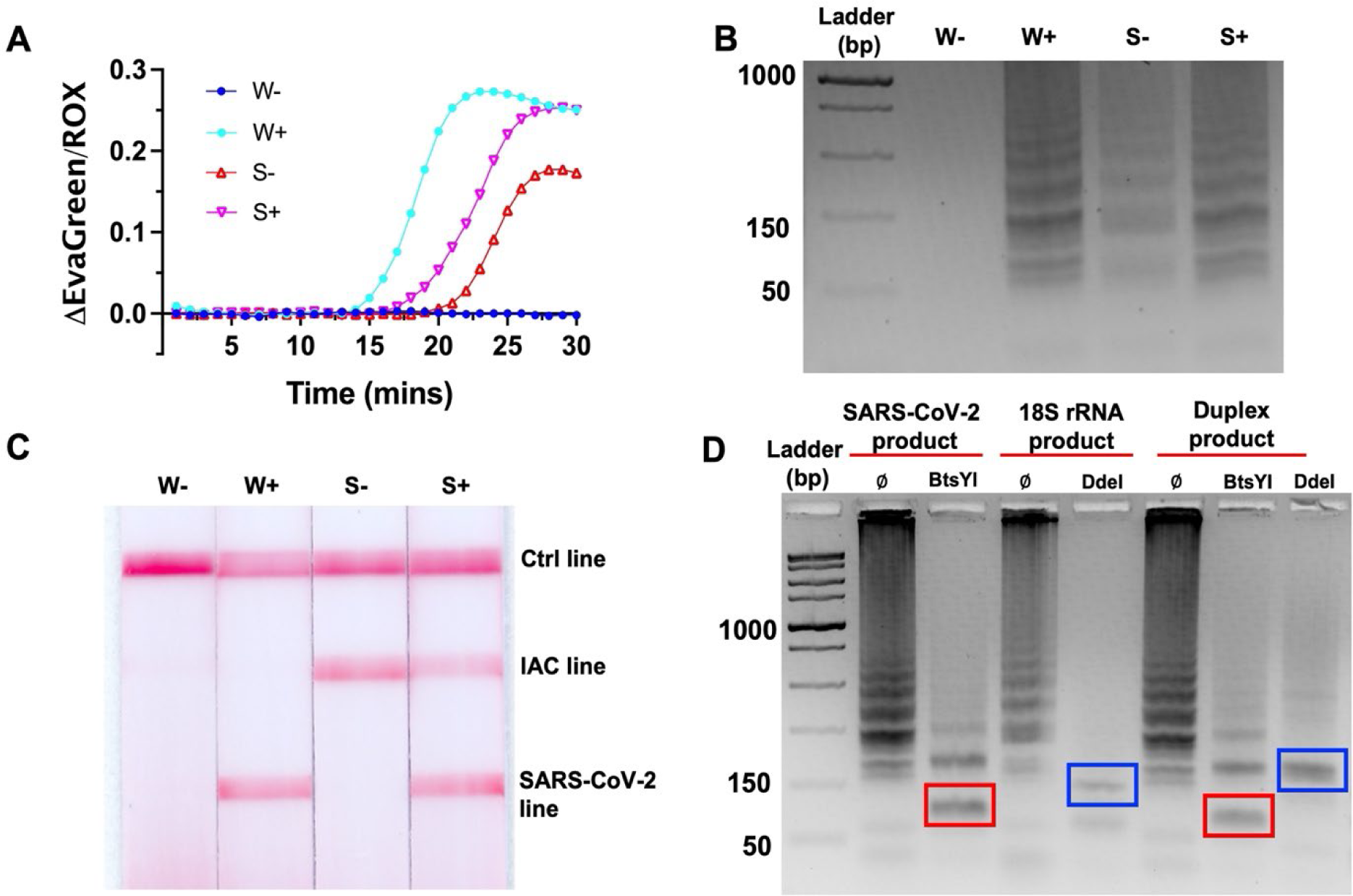
One-pot duplex RT-LAMP of SARS-CoV-2 and human 18S rRNA. (A) Amplification plot of duplex RT-LAMP using different templates. (B) Gel electrophoresis and (C) corresponding LFIAs of optimized assay (n=3). Integration of 18S rRNA human sample control into duplex RT-LAMP assay demonstrating differentiation between saliva (with 18S rRNA) and water matrix (without 18S rRNA) and SARS-CoV-2 spiked into each matrix. W- and W+ represent water with and without SARS-CoV-2, respectively. S- and S+ represent saliva sample with and without SARS-CoV-2, respectively. (D)

Restriction enzyme digestion of duplex RT-LAMP of SARS-CoV-2 and human 18S rRNA visualized on the gel. Red boxes indicate BstYI digested amplicon with the characteristic band at 110 bp and blue boxes indicate DdeI digested amplicon the characteristic band at 150 bp. Ø indicate non-digested amplicons.

### 3.4 Validation of the duplex RT-LAMP products by restriction enzyme digestion

The amplification products of the two primer sets in the duplex RT-LAMP were also confirmed by restriction enzyme digestion by using two specific restriction enzymes: BtsYI, which targets the product of SARS-CoV-2 primers, and Ddel, which targets the IAC primers (Figure 3D). The digested and non-digested amplicons of SARS-CoV-2, 18S rRNA, and duplex products were shown in Figure 3D. For SARS-CoV-2 product (SARS-CoV-2 spiked water), the anticipated 110 bp band was observed after BtsYI digestion as seen in lane 2 on the gel. This band did not show up in non-digested SARS-CoV-2 product in lane 1. In case of 18S rRNA product from saliva without SARS-CoV-2, the characteristic band at approximately 150 bp was observed when the amplicons were digested with DdeI as shown in lane 4. In lane 3 of non-digested product, the band at 150 bp was absent. Once the duplex RT-LAMP products from SARS-CoV-2 spiked saliva were digested with BtsYI and DdeI, the characteristic band at 110 bp and 150 bp were visible on the gel in lane 6 and lane 7, respectively, along with smear bands of undigested amplicons. This experiment confirms that the duplex RT-LAMP assay could amplify two targets in the one-pot reaction without cross-reactivity.

### 3.5 Analytical sensitivity of duplex RT-LAMP assay of SARS-CoV-2 and IAC

The LOD of the duplex RT-LAMP assay was determined by detecting different concentrations of SARS-CoV-2 spiked into saliva, as was performed in the singleplex SARS-CoV-2 RT-LAMP assay. As seen in Figure 4A, the IAC line showed up in all samples due to the presence of intact human 18S RNA in saliva. The leftmost strip of no viral particles represented a valid negative result. The strips of concentrations ranging from 100-5000 viral copies/µL demonstrated 3 lines: flow control, IAC, and SARS-CoV-2 test lines. Figure 4B demonstrates that there is a statistically significantly difference in the test band of duplex RT-LAMP products ranging from 100-5000 viral copies/µL compared to no template, as confirmed by a one-way ANOVA with Dunnett’s *post hoc* analysis. As a result, the duplex RT-LAMP exhibited a LOD of 100 viral copies/µL saliva comparable to single plex SARS-CoV-2 RT-LAMP. With prior heat-inactivation of saliva samples, we could bring the LOD down to 50 viral copies/µL (Figure S5). Moreover, we found that the IAC line intensity demonstrated a statistically significant decrease as the concentration of SARS-CoV-2 in the sample was increased. On the other hand, the SARS-CoV-2 test line intensity increased in a concentration-dependent manner. Importantly, while the IAC line remained visible in all tests, we hypothesize that in cases of exceptionally high viral loads of SARS-CoV-2, the viral target could outcompete the IAC in samples. This would lead to the absence of IAC at excess SARS-CoV-2 concentrations. In this case, users would be informed to interpret any result with a SARS-CoV-2 line as a positive result. The LODs of SARS-CoV-2 RT-LAMP assays can vary depending on several factors such as the type of assay used, the quality of the sample, and the target genes selected for amplification. As compared to several studies that have reported LODs of SARS-Cov-2 ranging from 10 to 1000 viral copies/µL [40–43], our duplex RT-LAMP could detect both targets without compromising the sensitivity of SARS-CoV-2 primers.

**Figure 4.**
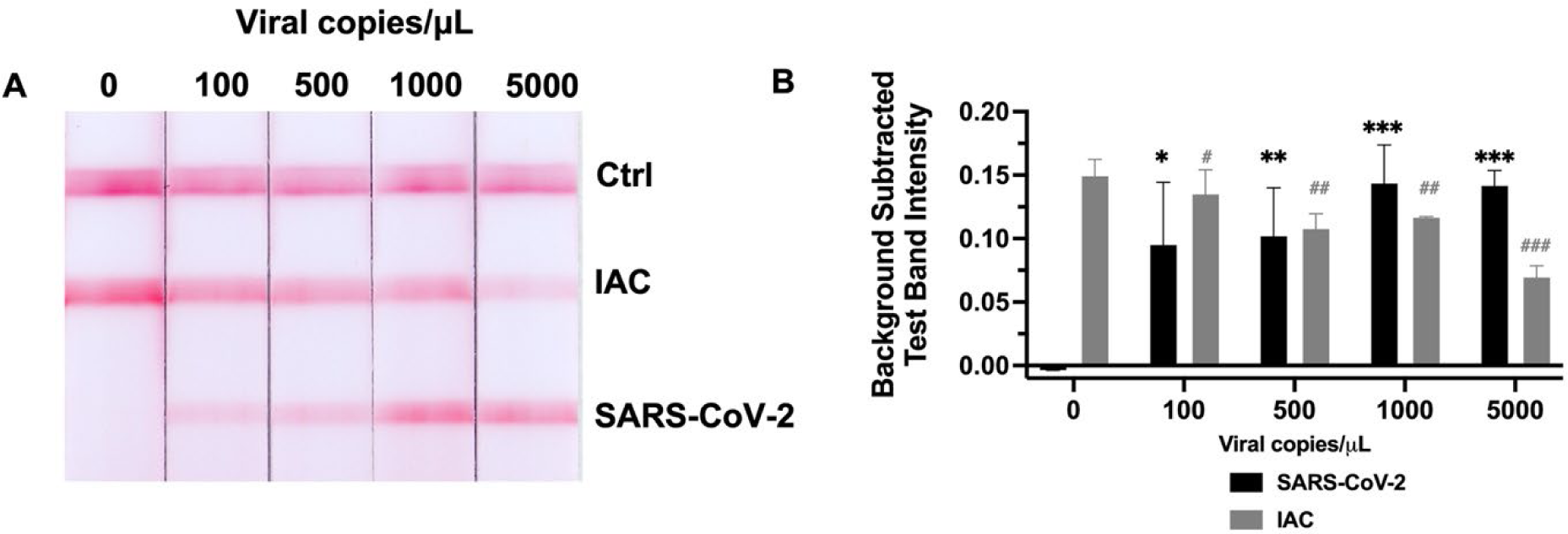
Analytical sensitivity of duplex RT-LAMP assay of SARS-CoV-2 and IAC in saliva visualized on (A) LFIA and (B) corresponding test band intensity analysis. n = 3; */# indicates *p* value ≤ 0.05; **/## indicates p ≤ 0.01; ***/### indicates *p* value ≤ 0.001; compared to equivalent bands at 0 viral copies/μL.

### 3.6 Clinical sample validation

Clinical sample validation is crucial for the successful implementation of diagnostic assays into clinical practice. It ensures that the assay is accurate, reliable, and can be used to make informed decisions about patient care. To evaluate clinical performance of our assay, we tested 30 deidentified clinical samples received from the Indiana Biobank. Since we intend to implement this assay at point-of-care sites, we bypassed any RNA extraction steps and validated the duplex assay by using non-extracted samples. The duplex RT-LAMP assays were run in triplicates for each sample and the results were interpreted on triple-line LFIAs. All LFIA results and quantified test line intensities are shown in Figure S6, Figure S7, and Table S2. We found that the IAC lines were observed in all samples although the IAC lines of non-extracted sample ID 10 were faint. Therefore, we ran the assay again with extracted samples and found that the IAC lines showed up clearly from this extracted sample ID 10 (Figure S5.) and Ct value of 22 obtained from RT-PCR indicated very high viral load [44]. Saliva sample ID 19 was highly viscous. Therefore, we diluted this sample with the nuclease-free water in a 1:1 ratio before adding to the reaction. Sample dilution is one of sample preparation methods for diagnostic tests that has been shown to help reduce inhibitory factors and viscosity of sample matrices [42].

The RT-PCR assay of all 30 extracted samples was analyzed as a reference method. The Ct values were used as a parameter for result interpretation according to manufacture (Table S3). The clinical sensitivity and specificity of the RT-PCR kit were shown in Table 2. The results suggest that our duplex RT-LAMP assay with non-extracted samples correctly identified 20 of the 21 RT-PCR–positive samples and accurately detected all 9 SARS-CoV-2 negative specimens. Only non-extracted sample ID 6 was identified as negative while the RT-PCR identified as positive. Sample ID 6 had an RT-PCR Ct value of 33, which is considered to be a very low viral load [45]. When RNA was extracted from sample ID 6, the RT-LAMP assay correctly identified the sample as positive. In general, the viral load of SARS-CoV-2 in saliva may vary throughout the different stages of infection. The study conducted by Juanola-Falgarona et al. shows that the viral load of SARS-CoV-2 in clinical samples exhibited linear relationship with Ct value. The lowest concentration of 1.00E+02 copies/mL corresponded to Ct value of 34.9 ± 3, while the highest concentration of 1.00E+06 corresponded to Ct value of 23.4 ± 0.7 [46].

**Table 2.** Calculation of sensitivity and specificity for the Duplex RT-LAMP.

|  |  | RT-qPCR |  |
| --- | --- | --- | --- |
|  |  | Positive | negative |
| Duplex RT-LAMP | positive | 20 (TP) | 0 (FP) |
|  | negative | 1 (FN) | 9 (TN) |

Sensitivity: 95%
Specificity: 100%
TP: True positive, FP: False positive, FN: False negative, TN: True negative

On non-extracted saliva samples, the duplex RT-LAMP achieves 95% clinical sensitivity, 100% clinical specificity, and 96% accuracy. These are considered well above the acceptable values according to the US Food and Drug Administration (FDA) guidance for SARS-CoV-2 diagnostic tests Emergency Use Authorization.

## 4. Conclusion

We successfully combined an IAC for clinically valid sample collection and assay function with a test for the SARS-CoV-2 virus from saliva using the one-pot duplex RT-LAMP. The test can detect both the target virus (SARS-CoV-2) and IAC in the same reaction without cross-reactivity. Without requiring RNA extraction, the duplex RT-LAMP assay was able to detect SARS-CoV-2 down to 100 copies/µL of saliva within 30 minutes, or 50 copies/μL with an additional heat inactivation step. The developed assay exhibited 95% clinical sensitivity and 100% specificity with accuracy of 96% on non-extracted saliva samples without heat inactivation. IACs are integral to ensure the accuracy and reliability and user confidence in molecular diagnostics in order to run them at home and at POC sites with minimally trained users. Additionally, both the specific 18S rRNA IAC and general knowledge of duplex RT-LAMP can be applied in similar manner to incorporate IACs into various other clinical sample matrices including blood, urine, and nasal swabs. This work is a promising step toward an integrated sample-to-answer POC device for respiratory infection detection at home or POC sites.

## CRediT authorship contribution statement

Navaporn Sritong: Conceptualization, Methodology, Investigation, Validation, Writing – original draft Winston Wei Ngo: Methodology, Investigation, Karin F. K. Ejendal: Methodology, Writing - review & editing. Jacqueline C. Linnes: Conceptualization, Writing-review & editing, Supervision, Funding acquisition.

## Data Availability

All data produced in the present work are contained in the manuscript

## Acknowledgement

This study received financial support from the Moore Inventor Fellows Award from the Gordon and Betty Moore Foundation Award # 9687 and from NIH National Institute on Drug Abuse award # DP2DA051910. We would like to recognize Dr. Mohit Verma, Assistant Professor in Agricultural and Biological Engineering, Purdue University, for providing information on SARS-CoV-2 primer sequences.

## Appendix A. Supplementary data

### Data availability

No data was used for the research described in the article.

---

**Table S1.**
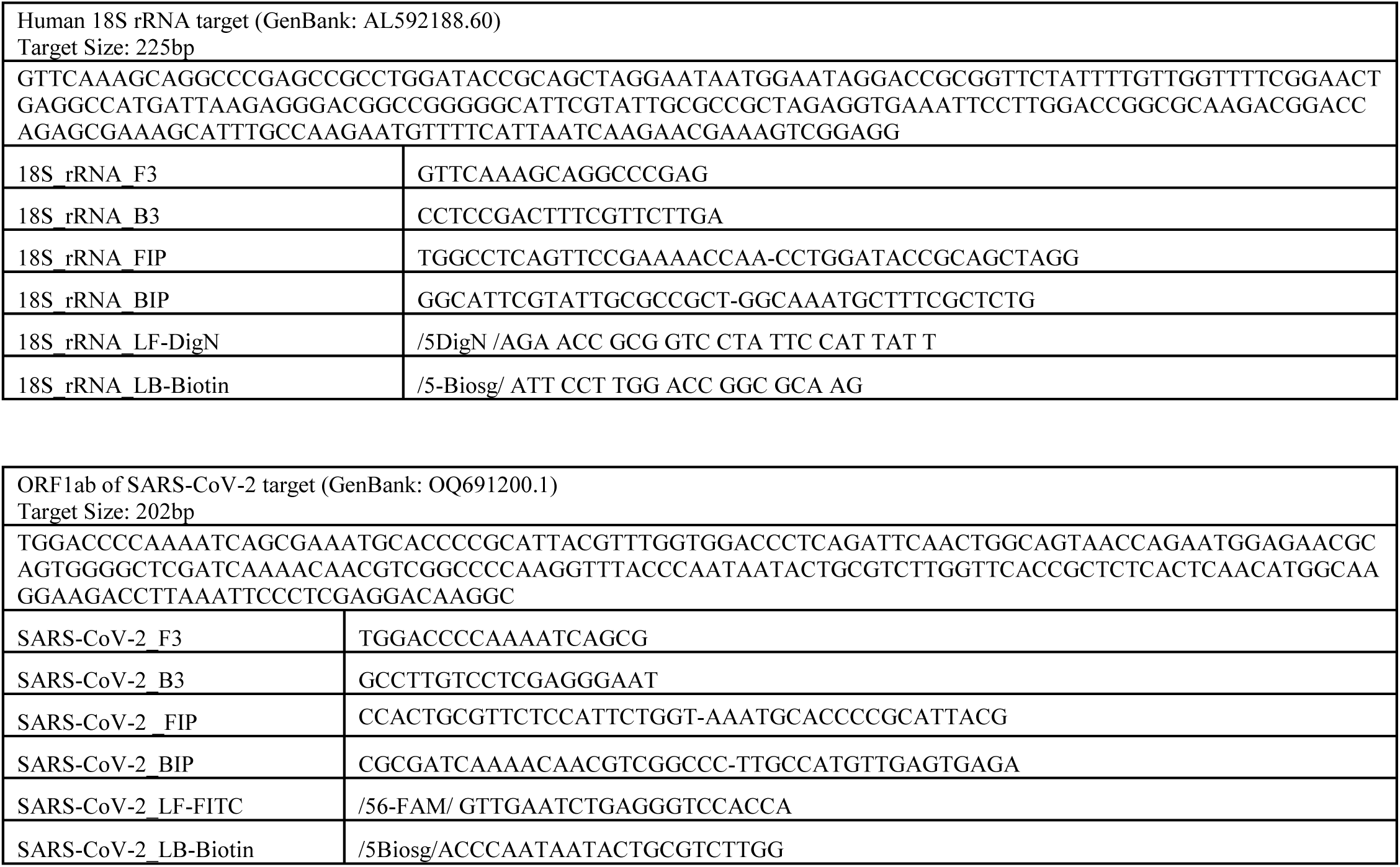
RT-LAMP primer sets targeting the 18S rRNA (IAC) and the ORF1ab gene (SARS-CoV-2 target)

**Table S2.**
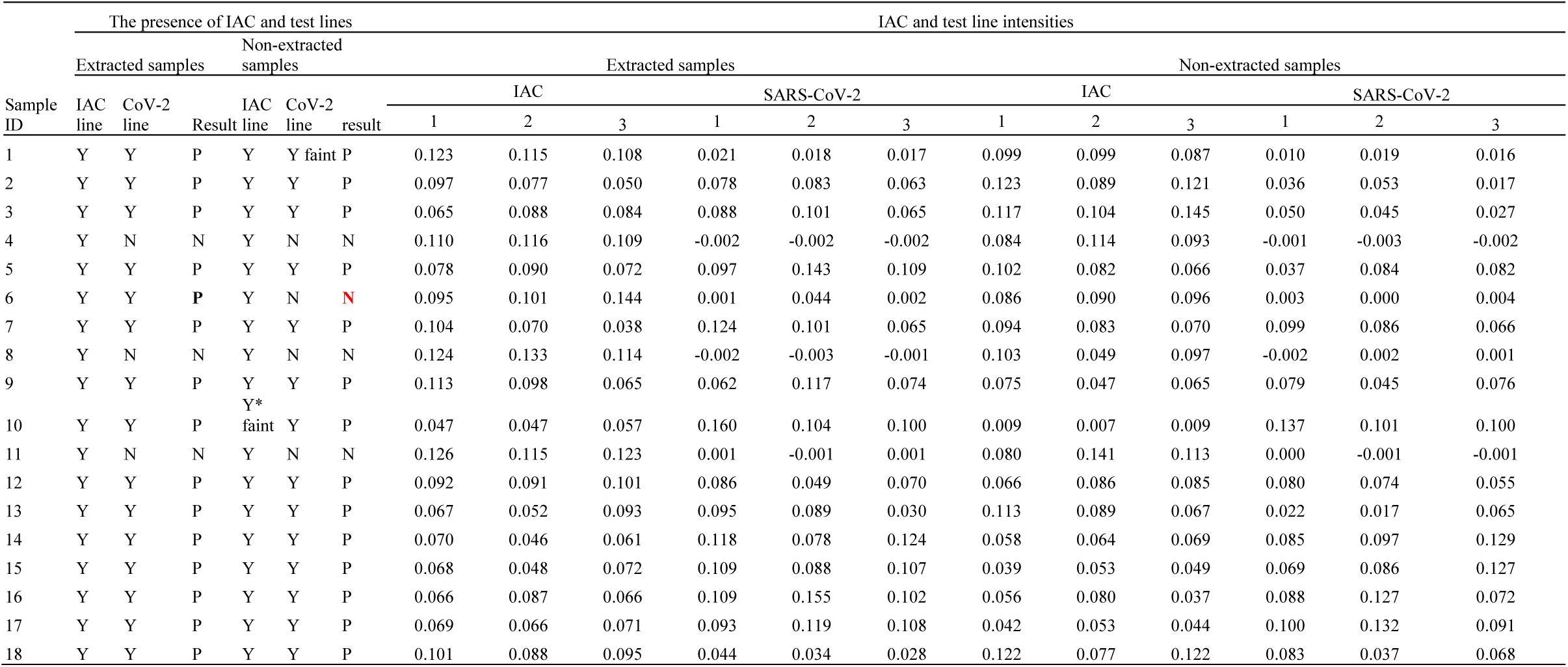
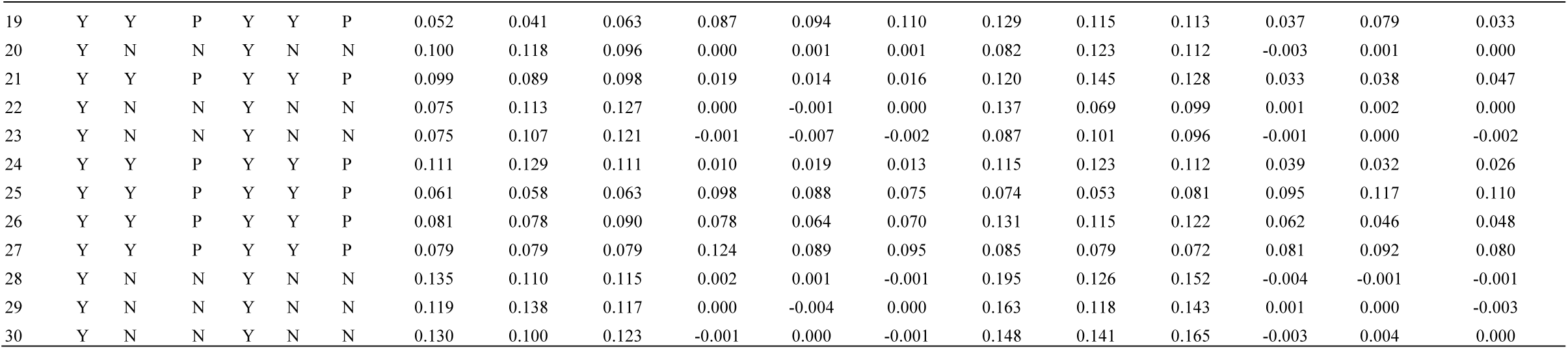
LFIA analysis of RT-LAMP performed on 30 clinical samples with and without extraction. IAC and test line intensities of each of 3 replicates of RT-LAMP assays are shown. Intensity of greater than 0.02 or greater is considered positive by eye [64]. P represents positive. N represents negative. Sample ID 6 was the only sample that indicated discordant results between extracted and non-extracted samples.

**Table S3.** Ct values of clinical sample ID 1-30. A positive result requires a sigmoidal amplification curve in the FAM channel with a Ct value of ≤37, and a sigmoidal amplification curve in the VIC/HEX channel with a Ct value of ≤35. A negative result is indicated by the absence of a sigmoidal amplification curve in the FAM channel with a Ct value of “0” or a sigmoidal amplification curve in the VIC/HEX channel with a Ct value of ≤35. P represents positive. N represents negative.

| Sample ID | Extracted samples |  |  |  | result |
| --- | --- | --- | --- | --- | --- |
|  | BGI qPCR |  |  |  |  |
|  | FAM ct |  | VIC ct |  |  |
| 1 | 30.372 | 33.506 | 21.911 | 23.348 | P |
| 2 | 35.325 | 35.133 | 28.100 | 28.778 | P |
| 3 | 24.379 | 24.372 | 23.274 | 24.505 | P |
| 4 | 0 | 0 | 24.562 | 24.940 | N |
| 5 | 29.733 | 30.525 | 20.450 | 20.599 | P |
| 6 | 33.643 | 33.833 | 26.562 | 26.226 | P |
| 7 | 26.822 | 25.953 | 25.323 | 23.809 | P |
| 8 | 37.132 | 37.489 | 24.375 | 25.993 | N |
| 9 | 30.965 | 29.877 | 22.641 | 22.267 | P |
| 10 | 22.319 | 22.117 | 24.264 | 24.843 | P |
| 11 | 0 | 0 | 25.485 | 25.638 | N |
| 12 | 31.001 | 31.219 | 22.754 | 22.738 | P |
| 13 | 32.311 | 25.038 | 34.340 | 25.641 | P |
| 14 | 27.356 | 27.021 | 29.878 | 29.464 | P |
| 15 | 25.753 | 25.065 | 24.059 | 22.999 | P |
| 16 | 22.972 | 23.046 | 23.004 | 22.862 | P |
| 17 | 23.417 | 21.674 | 23.398 | 21.527 | P |
| 18 | 32.618 | 33.312 | 30.102 | 29.607 | P |
| 19 | 21.441 | 21.083 | 24.302 | 24.517 | P |
| 20 | 0.000 | 0.000 | 24.115 | 24.192 | N |
| 21 | 32.384 | 32.205 | 25.081 | 25.211 | P |
| 22 | 0 | 0 | 29.103 | 27.581 | N |
| 23 | 0 | 0 | 20.637 | 24.378 | N |
| 24 | 33.581 | 35.209 | 21.984 | 21.707 | P |
| 25 | 26.267 | 26.419 | 23.427 | 23.957 | P |
| 26 | 30.643 | 24.207 | 32.105 | 25.253 | P |
| 27 | 29.981 | 30.303 | 26.780 | 27.682 | P |
| 28 | 0 | 0 | 30.147 | 30.456 | N |
| 29 | 0 | 0 | 27.319 | 25.731 | N |
| 30 | 0 | 0 | 32.767 | 32.927 | N |

**Figure S1.**
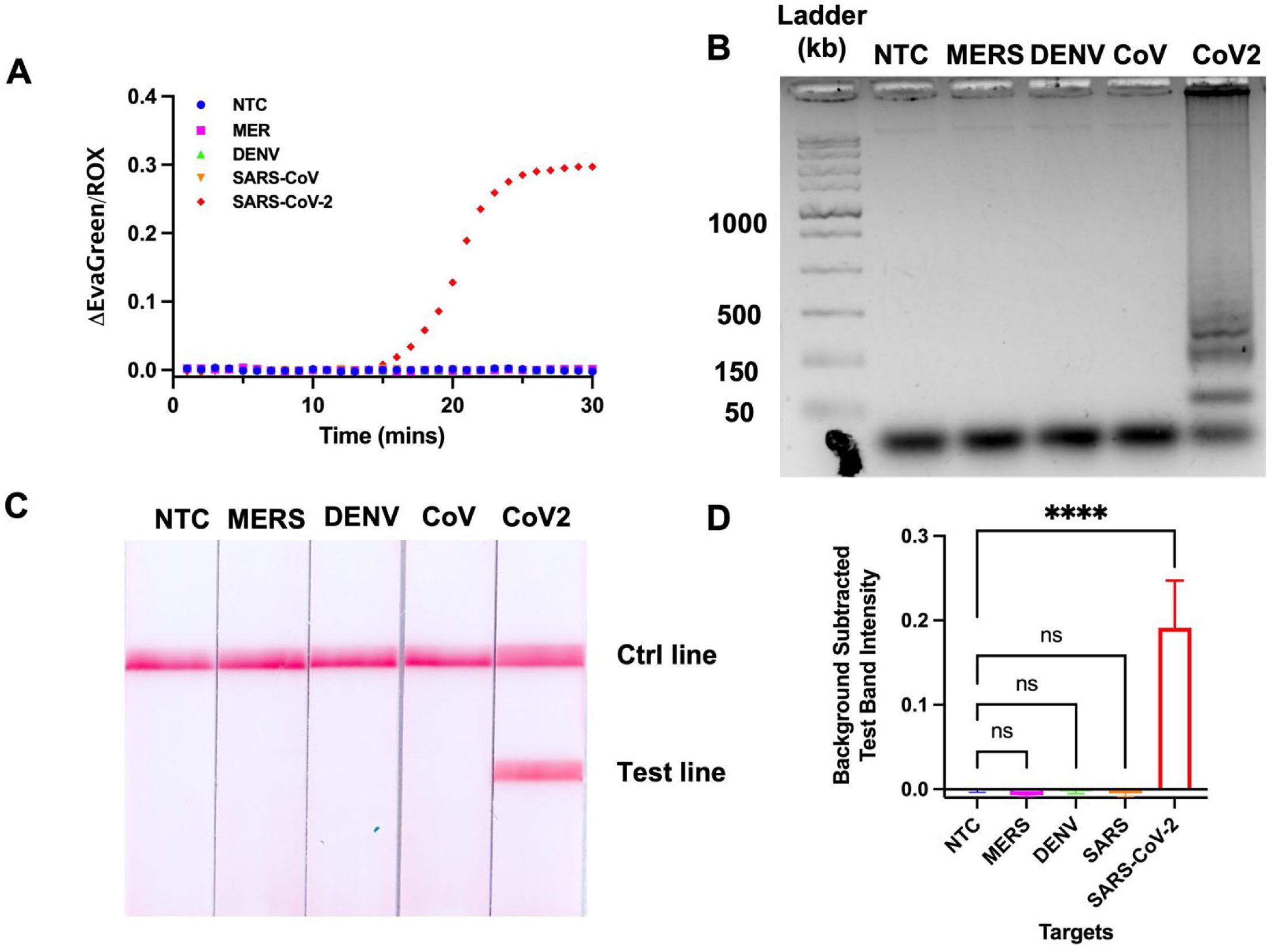
Analytical specificity of SARS-CoV-2 RT-LAMP assay against other viruses. (A) Amplification plot showed sigmoidal curve of SARS-CoV-2 template (B) Gel electrophoresis showed ladder pattern of amplified products of SARS-CoV-2 only. (C) The amplicons visualized on LFIA. Only reaction with SARS-CoV-2 generated test band. (D) corresponding test band intensity analysis. n = 3; **** indicates p ≤ 0.0001

**Figure S2.**
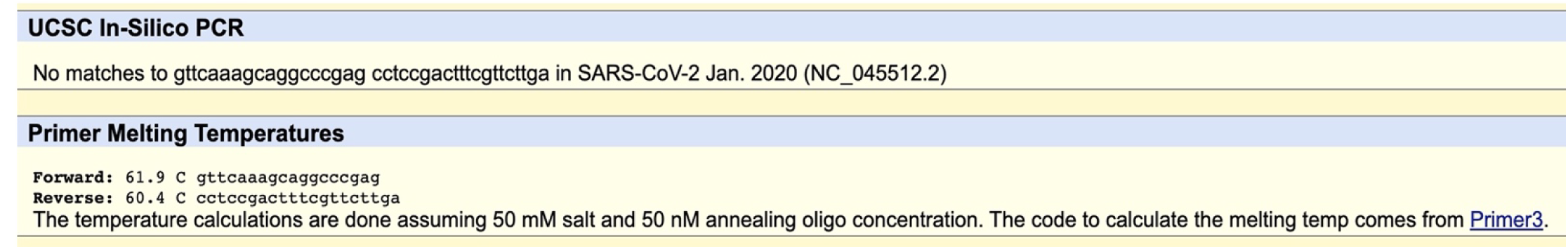
*In-silico* validation of 18S rRNA primer against SARS-CoV-2 genome.

**Figure S3.**
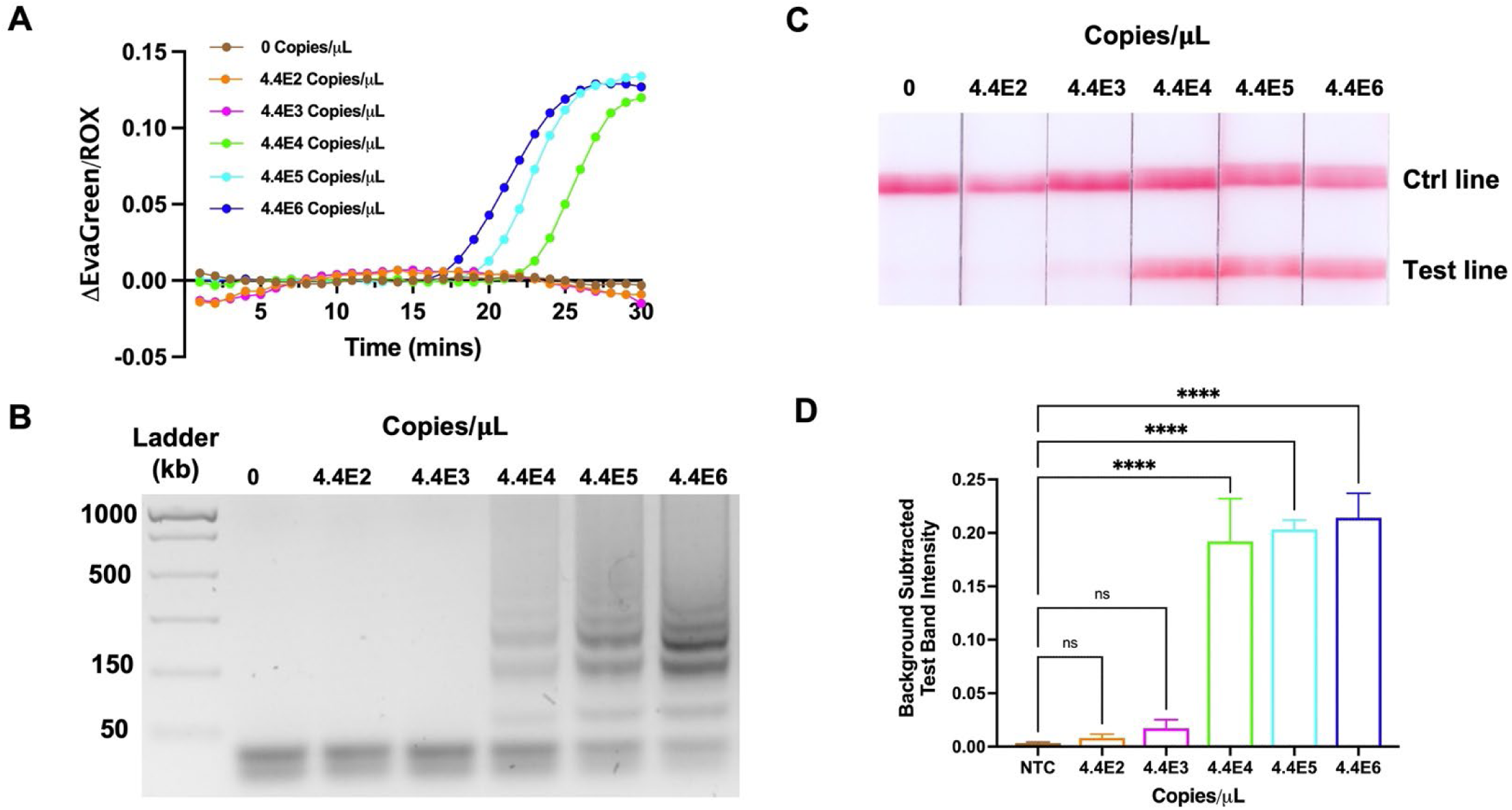
Analytical sensitivity 18S rRNA RT-LAMP assay. The amplification plot of 18S rRNA RT-LAMP using various concentration of human RNA control as templates (A). The corresponding gel electrophoresis analysis of 18S rRNA RT-LAMP amplicons (B). The LOD analysis on LFIA (C). The corresponding test band intensity analysis (D). n = 3; **** indicates p ≤ 0.0001.

**Figure S4.**
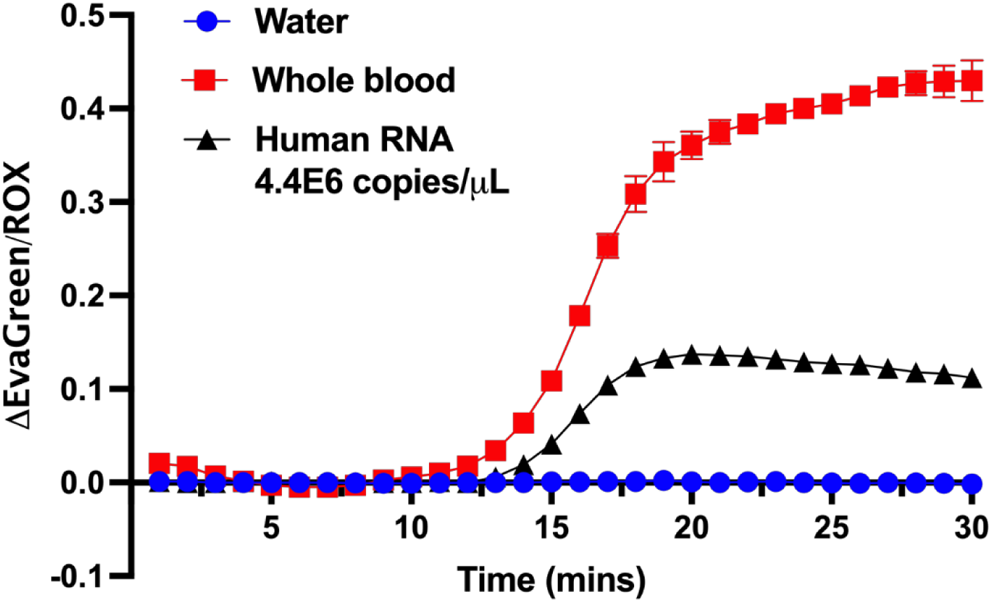
The usability of 18S rRNA as an IAC in blood sample. N = 2

**Figure S5.**
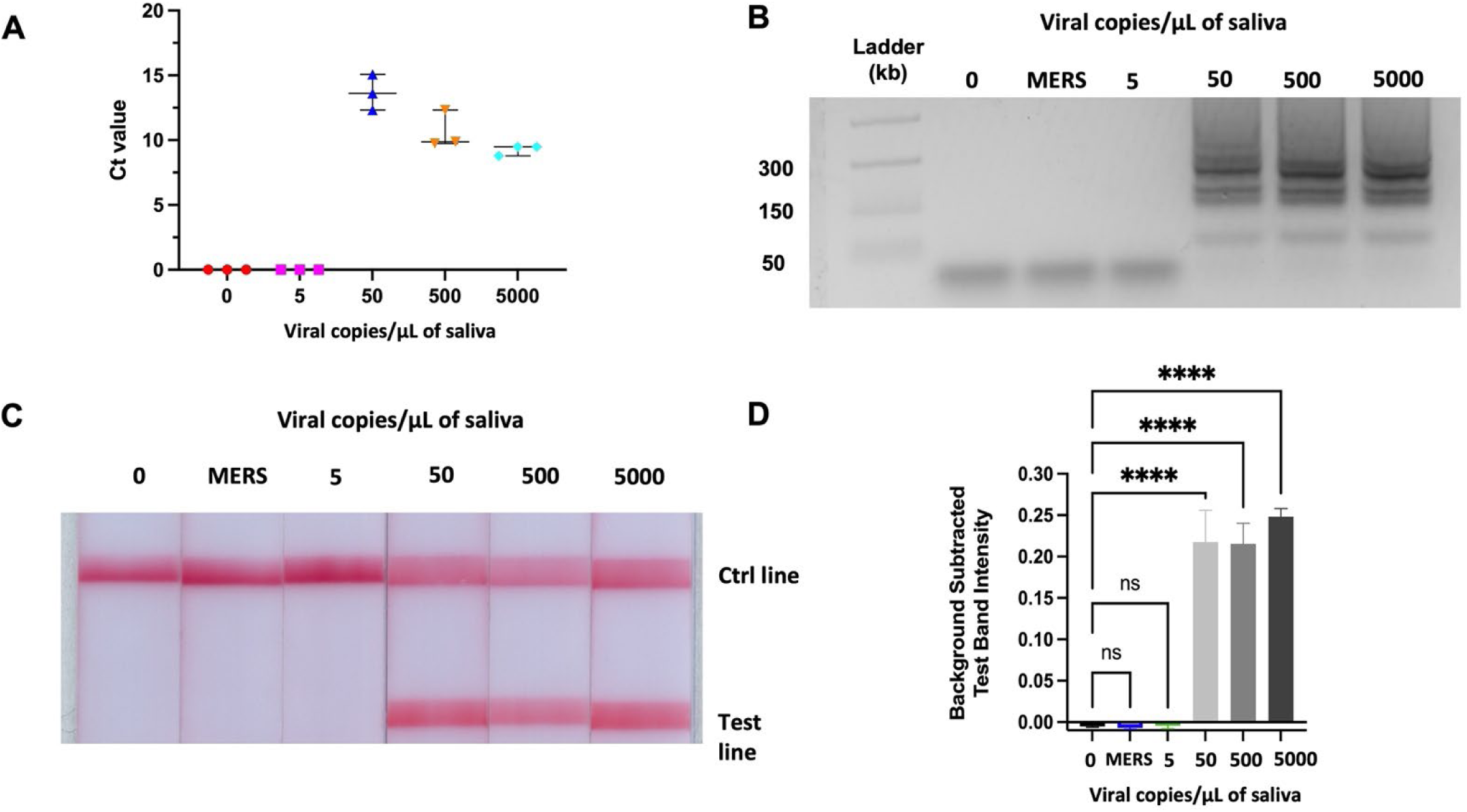
Analytical sensitivity of SARS-CoV-2 RT-LAMP assay in saliva with prior heat-treatment. RT-LAMP based detection of inactivated viral particles visualized on (A) gel electrophoresis (B) LFIA, and (C) corresponding test band intensity analysis. n = 3; **** indicates p ≤ 0.0001.

**Figure S6.**
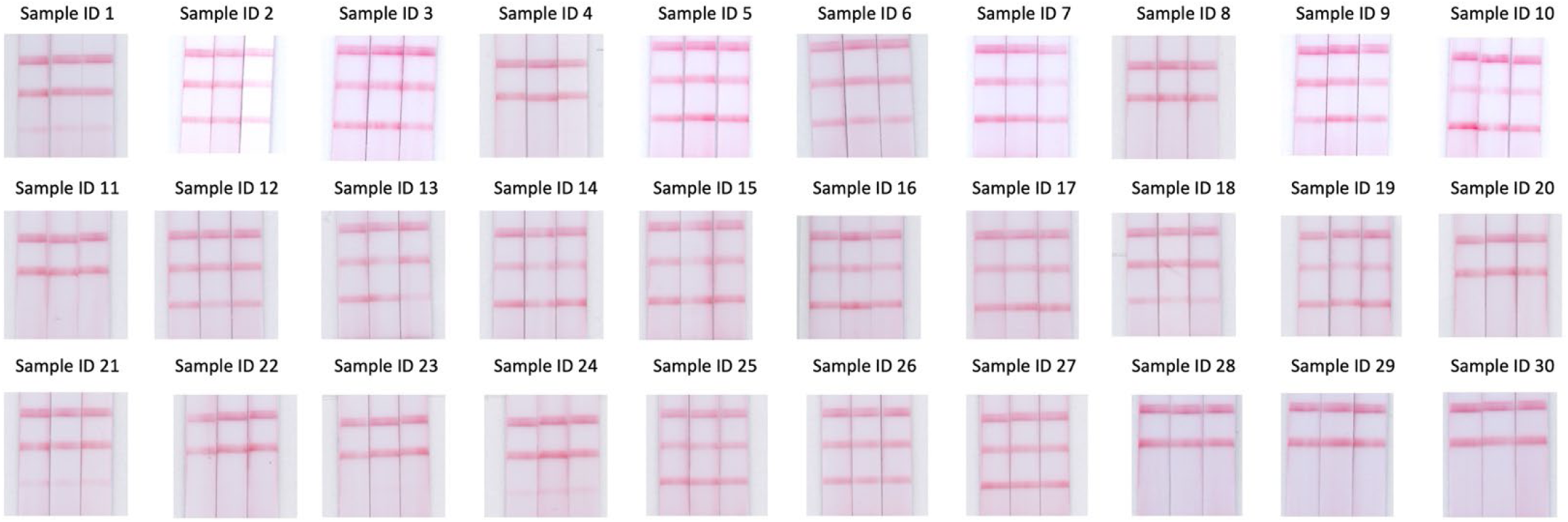
LFIA strips of extracted clinical samples, with the three lines representing “ctrl”, “IAC”, and test (from the top)

**Figure S7.**
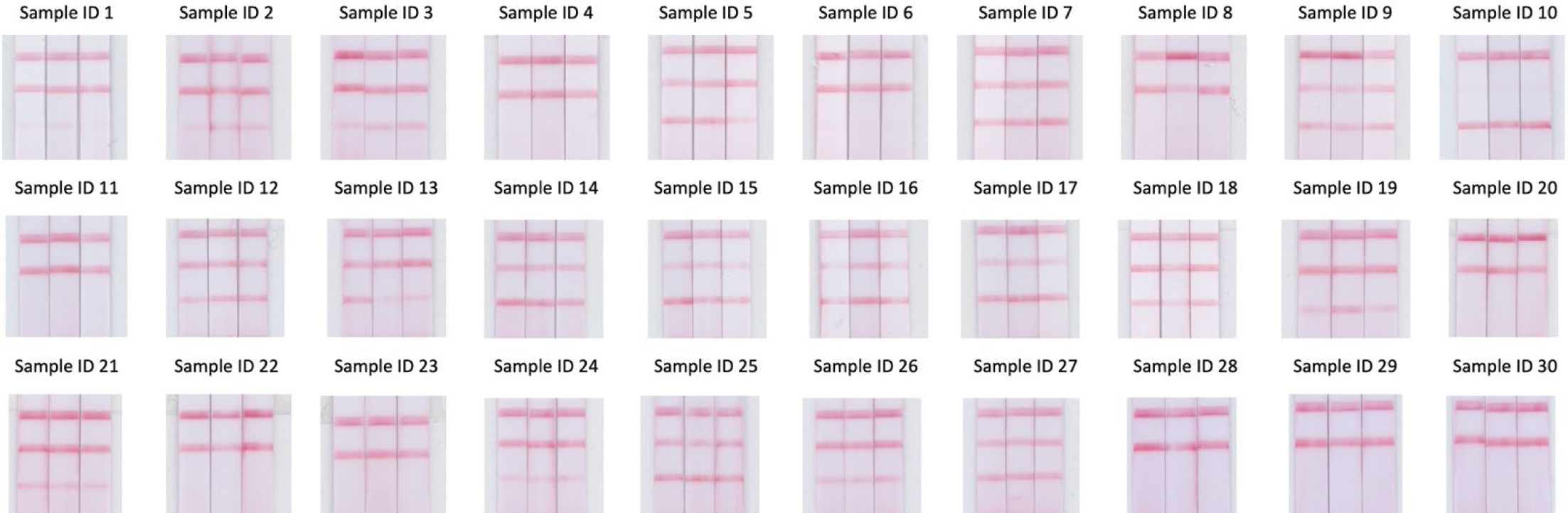
LFIA strips of non-extracted clinical samples

